# Benchmarking single-subject detectability of cognitive event-related potentials in healthy controls and minimally conscious state patients

**DOI:** 10.64898/2026.10.31.26366991

**Authors:** Yuri G. Pavlov, Melanie Boltzmann, Simone B. Jenner, Nick S. Menger, Maximilian Schwepcke, Steffen Gais, Nadezhda V. Pavlova, Damian Cruse, Jens D. Rollnik, Boris Kotchoubey

## Abstract

Event-related potentials (ERP) may reveal residual cognitive processing in disorders of consciousness, but clinical translation is limited because group-level effects do not guarantee reliable single-subject detection. Few ERP paradigms have been benchmarked in healthy participants and compared directly with patients in disorders of consciousness. We tested a hierarchical ERP battery in healthy controls and patients in minimally conscious state (MCS) to (1) estimate upper-bound single-subject detectability in healthy participants, (2) compare detection rates with those of MCS patients, and (3) evaluate prognostic value for recovery of consciousness. Forty-eight healthy controls and 26 patients in MCS completed six auditory paradigms: multifeature mismatch negativity (MMN), passive and active oddball P300, the subject’s own name, semantic oddball, and sentence N400. Effects were classified using blinded expert visual scoring, region-of-interest ANOVA, cluster-based permutation tests, and support vector machine decoding. Associations between binary ERP classifications and recovery were tested in 22 surviving patients at follow-up. In healthy controls, visual scoring identified MMN to at least one deviant and P300 in both oddball tasks in all 48 participants. Command following as demonstrated by task-related P300 enhancement between active and passive oddball conditions was detected in 81%. Command following was detected in 92% in the semantic oddball task. Sentence-congruity effects (N400 and/or P600) were identified in 69%, despite a strong group-level response, while the own-name effect was detected in only 35%. In patients, detection rates were lower: MMN was found in 27% for the short-duration deviant and 65% when any of five deviant types counted as positive; passive oddball P300 in 35% and active oddball P300 in 31%; sentence-congruity effects in 12%; active-versus-passive oddball modulation in 12%; and semantic oddball effects in 8%. Visual scoring and machine learning generally detected more effects than ANOVA and cluster-based testing, and agreement varied substantially across paradigms. Binary ERP classifications were not significantly associated with recovery (14 recovered, eight did not), although prognostic analyses were limited by sample size. Group-level ERP effects overstated single-subject detectability in healthy controls, whereas the absence of group-level effects in MCS obscured reliable ERP responses resembling those of healthy controls in a few individual patients. In MCS, ERP effects associated with semantic processing and command following were generally uncommon, and binary ERP indicators did not predict recovery in this sample. Our findings establish a benchmark for selecting and refining ERP paradigms and detection methods to assess residual cognition after severe brain injury.

## Introduction

Patients who survive severe acute brain injuries may evolve into chronic disorders of consciousness (DOC), most commonly unresponsive wakefulness syndrome (UWS) and minimally conscious state (MCS). MCS is defined by minimal but reproducible behavioral signs of self or environmental awareness^1^. Compared with UWS, patients in MCS have a substantially higher probability of regaining consciousness and functional communication^2^, making accurate prognostication particularly important.

Non-invasive measures such as EEG can reveal residual sensory, cognitive, and language processing not apparent at the bedside and may complement behavioral assessment in predicting outcome^3–5^. Although evidence is not yet strong enough to support routine clinical use^6–8^, several studies have reported that MMN, P300, and N400 are more often present in patients who later regain consciousness than in those who remain in UWS^9–11^. P300, particularly when elicited by subjectively meaningful or task-relevant stimuli^12,13^, as well as stronger late positive responses in self-referential paradigms using the subject’s own name^14,15^ have been associated with better outcomes. However, no single ERP paradigm currently provides sufficient accuracy for routine prognostic use, motivating the use of multi-paradigm approaches^3,4,8^.

ERP markers of higher-order processing may show stronger relationships with recovery than markers of low-level sensory change detection. We therefore adopted a hierarchical view of ERP paradigms that range from automatic sensory prediction violations to self-referential and semantic operations^16–18^. At the lower end of this hierarchy, mismatch negativity (MMN) and P3a elicited in passive oddball paradigms index automatic change detection and involuntary orienting. Higher in the hierarchy, P3b in active oddball tasks reflects instruction comprehension, maintenance of a task set, and sustained voluntary attention^3,19,20^. Larger P3b responses during active compared with passive conditions are typically interpreted as evidence of command following^21^. The subject’s own name elicits enhanced late positivity compared with other first names, probing recognition and autobiographical memory^14,15,22^. N400 paradigms probe sentence level semantic integration. N400 effects elicited by incongruent sentence endings have been observed in some patients with DOC^10,16^. The semantic oddball paradigm requires participants to count words from a specific semantic category and engages both command following and lexical semantic processing^23,24^.

The clinical usefulness of cognitive ERP paradigms is constrained by modest single-patient sensitivity and specificity^25^. In addition, clinical ERP protocols are not always systematically validated in healthy participants, so the baseline sensitivity of a given protocol under optimal conditions is often unknown. Even in healthy individuals, strong group-level effects do not guarantee single-subject detection, as demonstrated for N400^26,27^ and for P3b in active oddball tasks^28^. Kallionpää et al. (2019) found auditory N400 effects in 60–90% of participants depending on the analysis method, with conservative approaches missing effects evident on visual inspection. Paradigms intended for clinical use should therefore first demonstrate high single-subject sensitivity in healthy controls. Few studies have reported the detection rates in healthy volunteers and even fewer have compared these rates with those in DOC patients. We address this gap by testing healthy controls and MCS patients in six cognitive ERP paradigms.

Single-subject ERP detection methods include univariate statistics (ANOVA or t-tests) within predefined regions of interest^29,30^. Nonparametric cluster-based permutation tests^31^ reduce reliance on predefined electrodes and time windows while controlling familywise error. Multivariate machine learning methods exploit spatial and temporal patterns: support vector machines (SVM) and related classifiers can detect subtle but reliable differences between conditions and have been successfully used to decode covert command following in individual DOC patients^25^. Expert visual inspection remains a reference standard for EEG pattern recognition. In their systematic comparison of five single-subject N400 analysis methods, Kallionpää et al. (2019) found higher detection rates with visual scoring and some statistical approaches than with cluster-based tests, but no single method identified effects in every participant. We therefore compare expert visual scoring, region of interest ANOVA, cluster-based permutation tests, and machine learning with SVM.

We quantified single-subject detectability and prognostic value across a hierarchical ERP battery. First, we established baseline sensitivity for each paradigm and detection method in healthy adults, providing an upper bound for patient detection rates. Second, we applied the same battery and analysis pipeline to MCS patients and compared the presence of ERP components between groups. Third, we compared the four detection methods. Fourth, we entered binary single-subject classifications into multivariate logistic regression models to predict recovery of consciousness.

## Methods

### Participants

#### Healthy controls

Forty-eight healthy controls (33 female, 13 male, 2 other) participated in the study (mean age 31 years, SD = 11.5). Recruitment was conducted via mass emails at the University of Tübingen. Inclusion criteria were an age of 18-59 years and native German language proficiency. Exclusion criteria were any history of neurological or psychiatric disease and the use of any medication during the study. All participants provided written informed consent and received either 10 euro compensation or course credit.

#### Patients

Twenty-six patients with MCS were included. Data from the semantic oddball paradigm were missing for one patient, leaving 25 patients for that analysis. Mean age was 67.04 years (SD = 13.9). Mean time since brain injury to EEG recording was 67.04 days (SD = 24.7). See Table 1 for a description of the sample.

**Table 1:** Patient sample.

| ID | Sex | Age, y | MCS subtype | Etiology | Time since injury | CRS-R:<br>A/V/M/O/C/Ar | Outcome |
|---|---|---|---|---|---|---|---|
| **1** | M | 30-39 | + | Other | 116 | 3/2/0/1/1/2 | 2 |
| **2** | F | 50-59 | + | Vascular | 44 | 3/3/5/2/1/1 | 3 |
| **3** | F | 50-59 | + | Vascular | 83 | 3/3/5/2/1/2 | 3 |
| **4** | F | 60-69 | - | Vascular | 89 | 2/3/3/1/0/2 | 3 |
| **5** | F | 70-79 | + | Vascular | 51 | 3/3/5/1/0/2 | 3 |
| **6** | M | 70-79 | + | TBI | 42 | 1/2/2/1/1/2 | 3 |
| **7** | F | 50-59 | + | Vascular | 57 | 1/3/0/1/1/2 | 3 |
| **8** | M | 70-79 | - | TBI | 51 | 1/0/3/1/0/1 | 2 |
| **9** | M | 80-89 | + | Vascular | 55 | 3/2/4/1/0/2 | 2 |
| **10** | M | 80-89 | - | Vascular | 69 | 2/3/4/2/0/1 | 3 |
| **11** | M | 80-89 | + | Other | 96 | 4/5/4/2/1/2 | 3 |
| **12** | F | 80-89 | + | TBI | 55 | 3/2/4/1/0/2 | 3 |
| **13** | M | 70-79 | - | Vascular | 48 | 2/1/3/1/0/2 | 3 |
| **14** | M | 70-79 | + | TBI | 42 | 3/3/4/3/1/2 | 1 |
| **15** | F | 80-89 | - | Vascular | 68 | 2/3/2/1/0/2 | 1 |
| **16** | F | 70-79 | + | TBI | 40 | 1/3/4/2/1/2 | 2 |
| **17** | M | 60-69 | + | Vascular | 48 | 3/4/4/2/1/2 | 3 |
| **18** | M | 70-79 | - | Vascular | 58 | 2/3/4/1/0/1 | 2 |
| **19** | F | 50-59 | - | Anoxic | 65 | 1/2/2/2/0/2 | 3 |
| **20** | F | 40-49 | + | Vascular | 49 | 3/3/4/3/0/2 | 3 |
| **21** | M | 60-69 | + | Other | 64 | 2/4/4/1/1/2 | 1 |
| **22** | F | 40-49 | + | Vascular | 145 | 3/3/0/2/0/2 | 2 |
| **23** | M | 70-79 | + | TBI | 66 | 2/3/4/2/1/2 | 1 |
| **24** | F | 60-69 | - | Vascular | 82 | 1/3/4/1/0/1 | 3 |
| **25** | F | 60-69 | - | Vascular | 74 | 2/3/2/2/0/2 | 2 |
| **26** | M | 60-69 | + | Vascular | 86 | 3/2/2/1/0/2 | 2 |

*Note. CRS-R, Coma Recovery Scale–Revised; A, Auditory Function; V, Visual Function; M, Motor Function; O, Oromotor/Verbal Function; C, Communication; Ar, Arousal. Outcome: 1 = deceased; 2 = no recovery of consciousness (MCS or worse); 3 = recovery of consciousness (EMCS or better)*

Patients’ legal representatives received written information about the study and provided written consent prior to enrollment. They were informed that participation had no effect on medical treatment and that participation could be terminated at any time without any consequences.

Patients were admitted to the BDH-Klinic Hessisch Oldendorf (Institute for Neurorehabilitation Research, Hannover Medical School) and were included according to the following criteria: diagnosis of MCS, age between 18 and 89 years; time since injury of 1 to 6 months and stable circulation and respiration. Exclusion criteria were background EEG activity below 3 Hz, a history of depression, schizophrenia, drug abuse, sleep disorders, or any neurological disorders, and epileptic activity in EEG recordings.

The patients were assessed using the Coma Recovery Scale-Revised (CRS-R) 0-3 days before the EEG measurement^32^.

The study was approved by the local Ethics Committee of the Faculty of Medicine of the University of Tübingen and the Ethics Committee of Hannover Medical School (No. 7485).

### Procedure

The order of paradigms was fixed within each group. In healthy controls, paradigms were presented in the following order: N400, passive oddball, active oddball, mismatch negativity (MMN), subject’s own name (SON), semantic oddball. In patients, the order was: SON, passive oddball, active oddball, N400, MMN, semantic oddball. See Figure 1 for a graphical representation of each paradigm.

**Figure 1:**
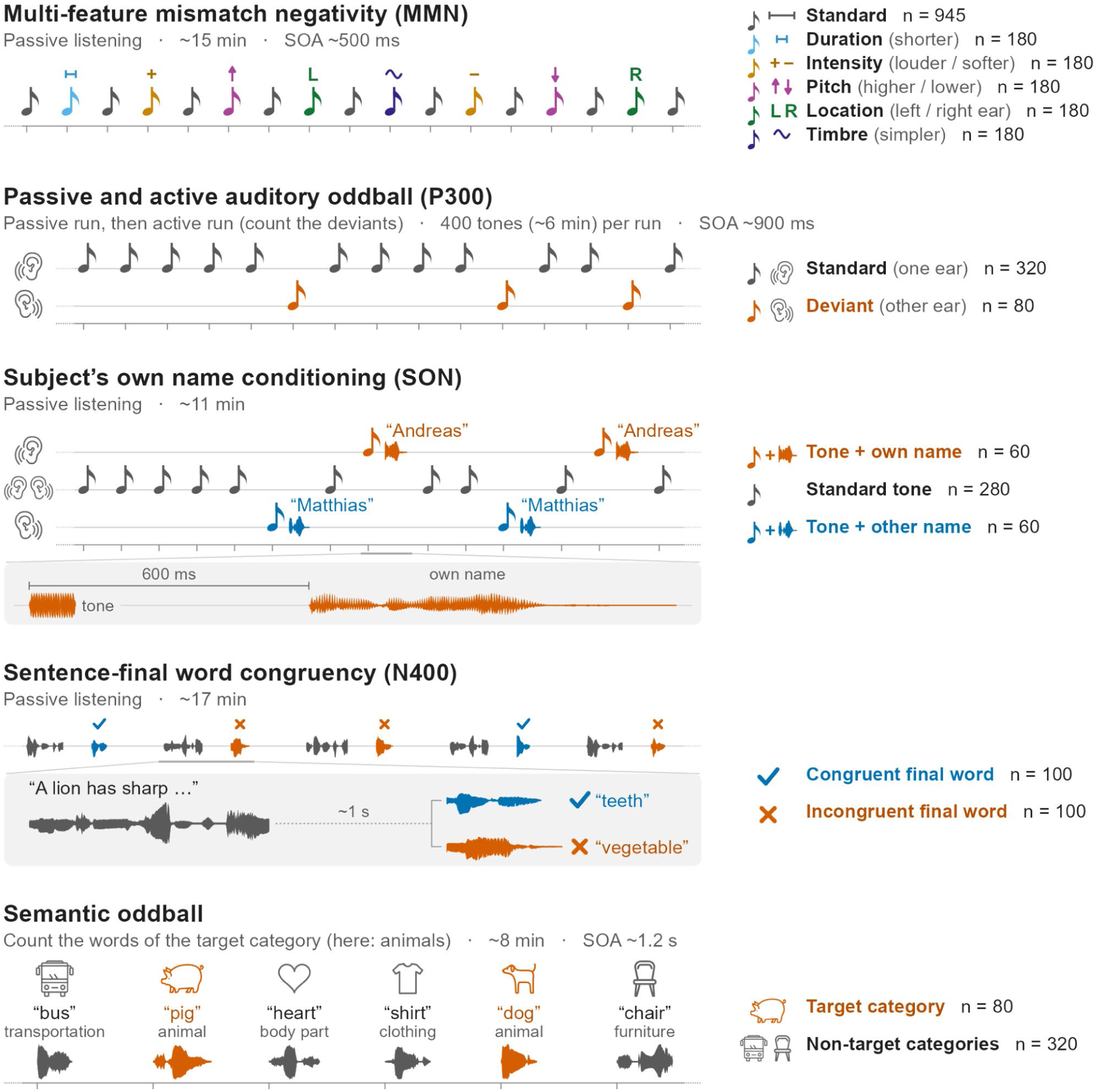
Visual depiction of the experimental paradigms.

**Figure 2:**
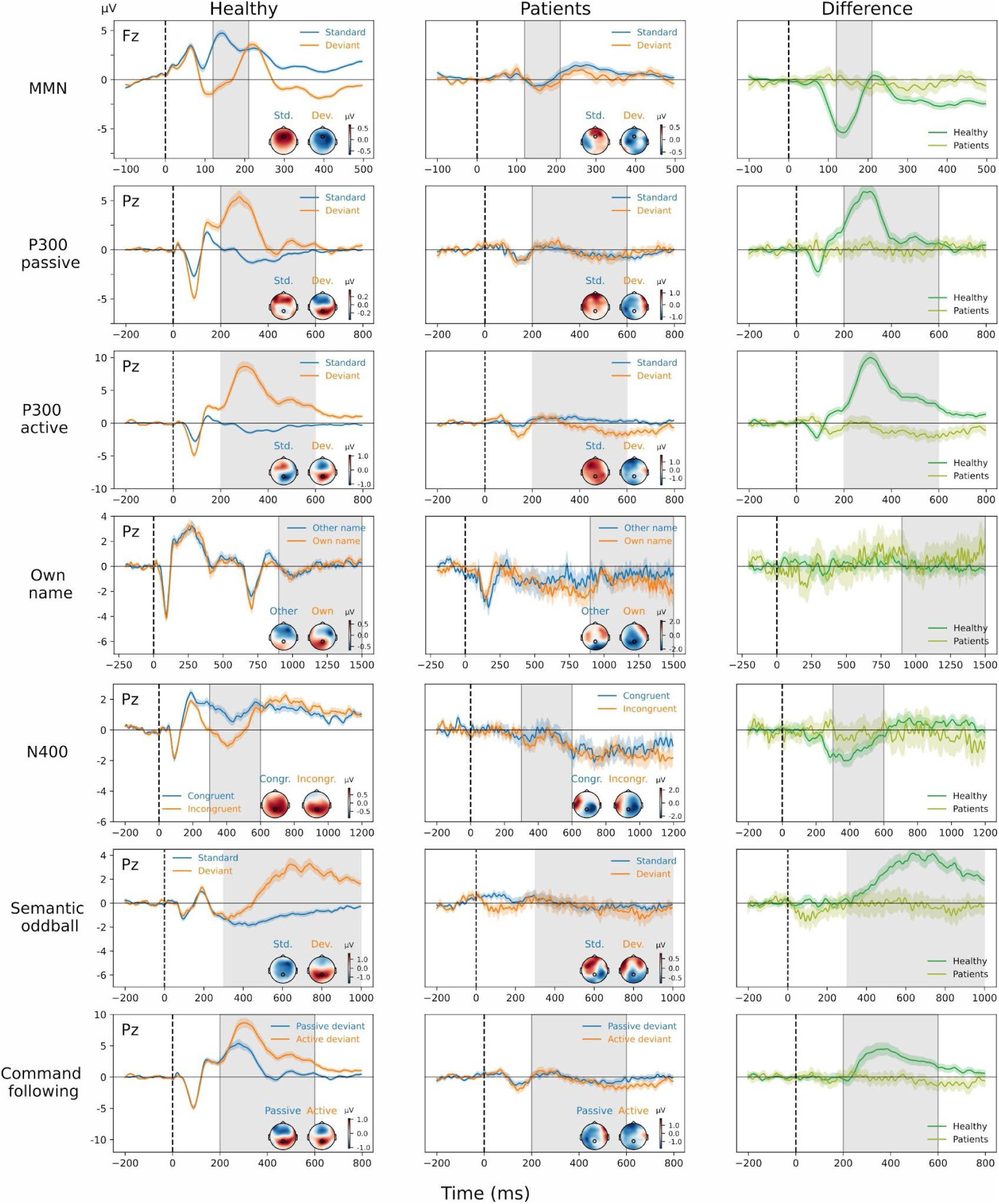
Grand average ERPs in each paradigm and both groups. Grey shading marks the time window for ANOVAs, cluster-based permutation tests, and SVM. The displayed waveforms are taken from the marked channels. For MMN, only standards and one deviant type are shown (plus the difference) to avoid overcrowding. All deviant waveforms for MMN are provided in the Supplementary.

All stimuli were delivered auditorily via pneumatic earphones (3M E-A-RTONE). Participants in both groups were seated in a comfortable chair throughout the experiment. To better mimic hospital conditions, healthy participants were not instructed to keep their eyes open or closed, but they were asked to avoid moving unnecessarily during the tasks.

### Experimental paradigms

#### Multifeature Mismatch negativity

Mismatch negativity (MMN) was assessed using a multifeature paradigm, also known as the Optimum-1 paradigm^33,34^. Frequent harmonic standard tones were interspersed with multiple deviant types that differed in acoustic features or spatial presentation, allowing efficient assessment of several auditory discrimination processes within one sequence.

The task comprised three blocks of 615 tones. Each block started with 15 standards and then alternated between 300 standards and 300 deviants. In total, 1845 stimuli were presented, including 945 standards and 900 deviants (180 of each of the five deviant types; intensity, pitch and location deviants were split equally between their two variants, 90 each). Deviants differed from standards in only one dimension and included: (1) duration (shorter), (2) intensity (louder or softer), (3) pitch (higher or lower), (4) location (left-only or right-only presentation), and (5) timbre (simplified spectrum).

Standard stimuli were harmonic tones with a fundamental frequency (f1) randomly drawn between 300 and 500 Hz. Each f1 was used to construct a harmonic complex with components at f1, f1*2, f1*4, f1*8, and f1*16. Standards were 75 ms long with 5 ms fade-in and 5 ms fade-out and were presented at 60 dB SPL.

Pitch deviants increased the fundamental frequency by 50% (higher pitch) or halved it (lower pitch). Intensity deviants were presented at approximately 70 dB (louder) or 45 dB (softer). Duration deviants were shortened to 25 ms. Location deviants were presented monaurally to the left or right ear, while standards were binaural. Timbre deviants retained only the fundamental frequency (f1) and removed higher harmonics.

Stimulus onset asynchrony (SOA) varied between 475 and 525 ms. The paradigm lasted approximately 15 minutes.

#### Passive and active oddball

Participants completed passive and active oddball tasks using the same stimulus parameters. In total, 400 stimuli were delivered (320 standards, 80 deviants). Each stimulus lasted 100 ms with 5 ms fade-in and 5 ms fade-out. SOA was 850-950 ms. Stimuli were delivered monaurally so that all standard tones were played to one ear and deviant tones to the other ear.

Tones were harmonic complexes with five components (f1, 2*f1, 4*f1, 8*f1, 16*f1). For each participant, the fundamental frequency (f1) was randomly selected from a set of four values for each stimulus type (standards: 300, 400, 440, 500 Hz; deviants: 95, 120, 150, 160 Hz). The first five stimuli were always standards. The remaining sequence was pseudorandomized with the constraint that deviants could not occur in immediate succession.

In the passive oddball task, participants were instructed to listen to the tones. The active oddball task started immediately after the passive task and used the same stimuli. Before the active task, participants were instructed to silently count the rare tones. Healthy participants reported counting an average of 79.3 ± 10.9 (mean ± SD) deviants.

Each oddball task lasted about 6 minutes.

#### Subject own name (SON) and name conditioning

Name conditioning is a variant of an oddball paradigm with an additional learning and recognition component (described in more detail in^35^). For brevity, we refer to this paradigm as the Subject’s Own Name (SON) paradigm.

A harmonic tone (conditioned stimulus, CS+) was paired with the participant’s own name (SON), and another tone (CS-) was paired with an unfamiliar name (other first name, OFN). A frequent standard tone was presented without any association. Tone-name pairing was counterbalanced across participants. OFNs were matched to the SON by the number of syllables. Name loudness was normalized to match the tones.

Conditioned tones and standards were 100 ms in duration (5 ms fade-in and fade-out) and consisted of harmonic tones created in a similar way to the P300 stimuli, but with different base frequencies and frequencies that were always different from those used in the other experiments. Stimuli were drawn from one of five sets of base frequencies (Hz): [162.5 247.5 495], [150 247.5 412.5], [97 150 247], [162.5 250 412.5], [195 300 495]. The standard tone was intermediate in pitch. SON and OFN tones were higher or lower in pitch relative to the standard.

OFN and SON were presented 600 ms after the paired tone onset, with a 500 ms silent interval between tone offset and name onset. After standards, SOA was 1300-1400 ms. After CS+ and CS-trials, the next stimulus followed the end of the name after 700-800 ms (SOA = name duration + 1300-1400 ms). Conditioned sounds (tone + SON and tone + OFN) were presented monaurally to opposite ears, with the side counterbalanced across participants.

The first five tones were always standards. Then 60 tone+SON and 60 tone+OFN stimuli were interspersed among the remaining 275 standards. The experiment lasted about 11 minutes.

#### N400

The N400 paradigm included 200 sentences, with 100 semantically congruent and 100 incongruent cloze endings. After the end of each sentence, ISI was 950-1050 ms. After the cloze word, ISI was 1950-2050 ms. Participants were instructed to listen to the sentences. Instructions were delivered in a prerecorded female voice. The experiment lasted about 17 minutes.

#### Semantic oddball

Participants completed a semantic oddball task that had been conceptualized previously, a version of which was validated in healthy controls^23^ and DOC patients^30^.

A total of 400 spoken words were presented from five semantic categories: body parts, transportation, clothing, animals, and furniture. Twenty-five distinct words were selected (five per category) and each word was presented 16 times, yielding 400 presentations. Of these, 320 were non-target (ignored) and 80 were target (counted) words from a randomly selected semantic category. SOA varied between 1150 and 1250 ms. Participants were instructed to silently count words from the target category. Instructions were delivered in a prerecorded female voice.

Healthy participants counted an average of 71.7 ± 13.1 (mean ± SD) words. Presentation order was randomized. The experiment lasted approximately 8 minutes.

#### Electroencephalography

EEG was recorded using two systems. Patients were recorded in the hospital with a 32-channel Brain Products LiveAmp system using BrainCap caps with built-in passive electrodes (Multitrodes). FCz served as the online reference and Fpz as ground. The sampling rate was 500 Hz. Healthy controls were recorded with a 64-channel Brain Products ActiCHamp system with active electrodes placed according to the extended 10-20 system. Cz served as the online reference and Fpz as ground. Sampling rate was 1000 Hz. Impedance was kept below 25 kOhm.

For subsequent analyses, recordings were harmonized by selecting the 32 electrodes that were present in both systems. Because passive and active electrodes can differ in their susceptibility to environmental noise, line noise was attenuated during preprocessing. In the hospital setting, line noise was not excessive and was effectively suppressed by low-pass filtering.

Preprocessing was identical for both groups and performed in EEGLAB^36^. Continuous data were filtered with a 0.5 Hz high-pass filter and a 45 Hz low-pass filter using the pop_eegfiltnew function with default parameters. A 0.5 Hz high-pass cutoff, rather than 0.1 Hz, was used to reduce prominent low-frequency artifacts in patient recordings, largely related to movement. Data were then downsampled to 250 Hz and rereferenced to the common average.

Independent component analysis (ICA) was computed using AMICA^37^. To improve ICA decomposition, a copy of the continuous data was high-pass filtered at 2 Hz for ICA training. The resulting ICA decomposition was then transferred to the dataset filtered between 0.5 to 45 Hz. Components reflecting eye movements and high-amplitude muscle activity were removed, as were components localized to a single electrode that could be clearly distinguished from physiological EEG activity.

ERP baseline correction used -200 to 0 ms for all paradigms except MMN (-100 to 0 ms). Epoch windows were as follows: N400, -200 to 1200 ms; passive and active oddball, -200 to 800 ms; SON, -200 to 1500 ms; MMN, -100 to 500 ms; semantic oddball, -200 to 1000 ms.

After epoching, remaining artifact-contaminated epochs were identified by visual inspection and discarded. Before statistical analysis, epoched data were re-referenced to the average of the mastoid electrodes.

#### Effect identification methods

For all automated procedures (ANOVA, permutation tests, machine learning), artifact-free trials were used for each participant. In the name conditioning paradigm, we compared responses to the names only because group-level findings in controls were weak or absent in the time windows corresponding to expected learning effects observed previously^38^. To assess command following in the active oddball paradigm, we compared P300 responses between active and passive oddball, expecting a larger P300 amplitude in the active task. In the N400 paradigm, both N400 and P600 ERPs were counted toward effect presence. Visual scoring used the same condition comparisons as the automated procedures.

#### Visual scoring

Two coauthors (YGP and BK) independently scored the presence of each ERP component of interest. To conceal group assignment, pooled patients and controls received new random IDs, retained across paradigms within each participant. Scorers reviewed ERP curves averaged by condition in multiple formats: (1) midline electrodes Fz, Cz, and Pz, (2) a multiplot showing all electrodes, and (3) global field power. Standard error of the mean was shown on the figures 1 and 3. After independent scoring, the scorers met multiple times to resolve disagreements and reach consensus. Initial agreement is shown in Figure 3.

**Figure 3:**
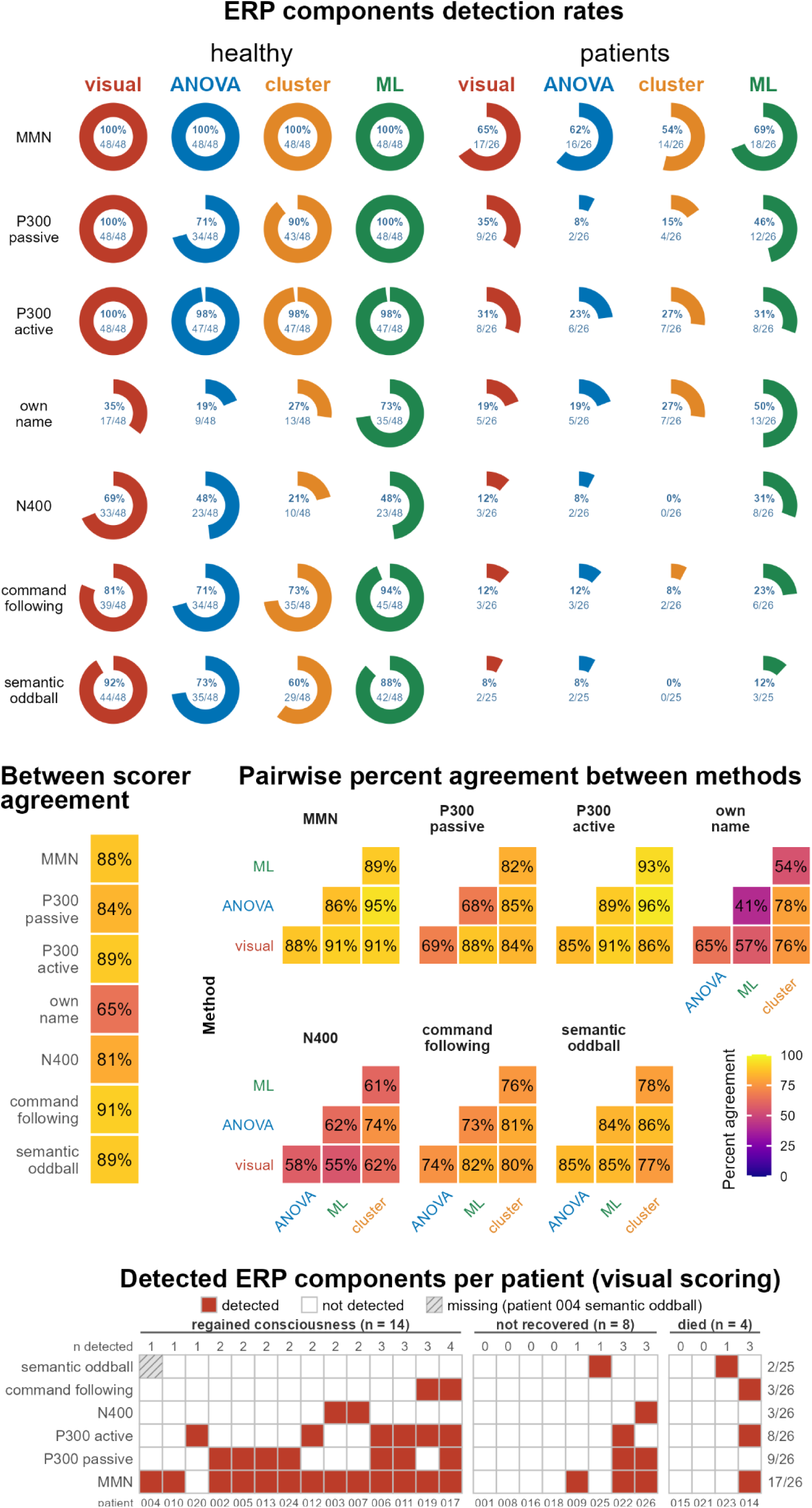
Comparison of single-subject ERP detection methods. Top: Donut charts show the percentage and number of healthy controls and patients in whom each ERP effect was detected, separately for each analysis method. Command following refers to the difference in P300 responses between active and passive oddball conditions. MMN detection refers to a response to at least one of the five deviant types. Middle: Agreement between the two visual scorers before consensus (left) and pairwise percentage agreement between detection methods (right). Bottom: Individual patient profiles showing ERP effects detected by visual scoring across paradigms. Missing data are indicated by hatching.

#### ANOVA

Following a previously established approach^30^, we used ANOVA in predefined time windows and channels (see Table 2 below).

**Table 2:** Time window and channel region of interest definition for ANOVA.

| Component | Time window, ms | Channels |
|---|---|---|
| P300 (active, passive) | 200-600 | Fz Cz Pz FCz |
| N400 | 300-600 | Cz C3 C4 CP1 CP2 P3 Pz P4 |
| P600 | 500-1200 | Cz C3 C4 CP1 CP2 P3 Pz P4 |
| Own name response | 900-1500 | Cz CP1 CP2 P3 Pz P4 C3 C4 |
| MMN | 120-210 | Fz FC1 FCz FC2 Cz |
| Late positivity in semantic oddball | 300-1000 | Cz CP1 CP2 P3 Pz P4 C3 C4 |

If either the main effect of Condition or the Condition x Channel interaction was significant (p < 0.05), we marked the component as present for that participant.

Analyses were implemented in R version 4.4.2.

#### Cluster-based permutation tests

Cluster-based permutation tests were run in FieldTrip^31^ with a minimum cluster size of 2 channels and 5000 permutations. The same time windows as in the ANOVA were used (except that N400 and P600 were tested together in a single 300–1200 ms window), but amplitudes were not averaged across time or channels, allowing the algorithm to detect significant clusters in time and space. Eighteen channels were used to reduce peripheral coverage while retaining most ANOVA channels plus additional channels: Fz, Cz, Pz, F3, F4, FC5, FC1, FCz, FC2, FC6, C3, C4, CP5, CP1, CP2, CP6, P3, and P4.

#### Machine learning with support vector machine

Single-subject decoding analyses were performed on ERP data using MATLAB’s fitcecoc function with SVM learners and one-vs-all error-correcting output codes. Analyses included 30 scalp channels (excluding mastoid electrodes) and used epoched data.

Decoding was performed separately for each participant and paradigm, using a time-window decoding approach: for a predefined time window of interest (TOI) specific to each paradigm (same as for ANOVA and permutation tests; see Table 2), features were constructed by concatenating voltages across all channels and all time samples within the TOI, producing a spatiotemporal feature vector for each trial.

To balance trial counts, each decoding run randomly sampled the minimum number of artifact-free trials per class (minTrials), where minTrials was the minimum available across included conditions for that participant. Each class contributed ceil(minTrials/2) trials to training and the remainder to testing. Accuracy, defined as the proportion of correctly classified held-out trials, was averaged across 10 runs with different random samples and splits.

Statistical significance of decoding accuracy was assessed with a label permutation test. For each of 1000 random permutations of trial labels, TOI decoding accuracy was averaged across ten random train-test splits to obtain a null distribution of decoding accuracy. The p-value was computed as (*k* + 1)/(*nPerm* + 1), where *k* is the number of permuted accuracies greater than or equal to the observed accuracy and *nPerm* = 1000.

### Group-level statistics

To describe group-level trends and estimate effect sizes, we used paired t-tests on mean amplitude averaged within the same time windows as those used for ANOVA. A single electrode was used for each effect, with Pz for all effects except MMN, where Fz was used.

### Outcome in the patients

Outcome data were obtained for 26 patients via a phone interview based on the GOS-E. Four patients were deceased at follow-up, 14 had regained consciousness (diagnosis EMCS or better), and 8 had not (MCS or worse). The average follow-up time from EEG recording to outcome assessment was 183.8 days (range 179-279 days).

## Results

### Group comparison

As manipulation checks and to estimate effect sizes, we tested standard effects for MMN, N400, and P300 in passive and active oddball tasks. We also tested the semantic oddball condition effect and the response to the participant’s own name, as a replication of our previous work.

In healthy controls, effects were strong across all paradigms except SON. Semantic oddball showed dz = 1.16, comparable to passive oddball with dz = 1.09 (see Table 3). Active oddball produced a larger effect size of 1.69. N400 also showed an effect size of similar magnitude, dz = 1.16. MMN had the largest effect size, with dz = 2.14 for shorter deviants, and deviant-specific effect sizes of 2.48 for intensity, 2.08 for pitch, 1.25 for location, and 1.98 for timbre. Unexpectedly, the response to the participant’s own name did not show a significant group-level effect.

**Table 3:** Results of the group comparison t-tests.

| ***Group*** | ***Experiment*** | ***t*** | ***p*** | ***dz*** |
|---|---|---|---|---|
| *healthy* | MMN | -14.821 | <0.001 | -2.14 |
|  | N400 | -8.025 | <0.001 | -1.16 |
|  | P300_active | 11.718 | <0.001 | 1.69 |
|  | P300_passive | 7.58 | <0.001 | 1.09 |
|  | SON | 0.322 | 0.749 | 0.05 |
|  | SemOddball | 8.049 | <0.001 | 1.16 |
| *patients* | MMN | -0.639 | 0.528 | -0.13 |
|  | N400 | -0.675 | 0.506 | -0.13 |
|  | P300_active | -2.357 | 0.027 | -0.46 |
|  | P300_passive | 0.047 | 0.963 | 0.01 |
|  | SON | -0.784 | 0.441 | -0.15 |
|  | SemOddball | -0.388 | 0.701 | -0.08 |

*Note. Negative t and dz values indicate the effect direction such that the difference wave is negative. MMN is reported for shorter deviants only.*

In patients, the only significant effect was observed for P300 in the active oddball task, but with the opposite sign, with a larger amplitude for standards than for deviants in the P300 time window.

### Scoring methods comparison

Cluster permutation tests yielded the lowest detection rates (Figure 3). In healthy controls, visual scoring achieved 100% detection for MMN and both P300 paradigms. Semantic oddball detection was also high, exceeding the command-following estimate from the active versus passive oddball comparison. Compared with visual scoring, machine learning showed similarly high MMN and P300 oddball detection, lower N400 detection, and numerically more command-following detections.

In patients, machine learning produced numerically higher detection rates than visual scoring.

### Interrater and intermethod agreement

Agreement was highest between cluster-based permutation tests and ANOVA, and lowest between ANOVA and machine learning (see Figure 3). Methods disagreed substantially in the SON paradigm. Agreement was also limited for passive oddball, falling below 70% for some method pairs. These pairs involved ANOVA, which detected the passive P300 in only 34 of 48 controls, whereas visual scoring and machine learning detected it in all controls. Agreement between methods for N400 was low (55–74% across method pairs).

For visual scoring, interrater agreement before consensus was above 80% for all paradigms indicating a good level of agreement^39,40^, except for the subject’s own name paradigm.

### Clinical outcome prediction

Excluding deceased patients, multiple logistic regression models tested whether binary single-subject classifications from each paradigm predicted recovery of consciousness (yes/no). No significant effects emerged, including when analyses were repeated separately for visual scoring, ANOVA, machine learning, and cluster permutation tests.

## Discussion

Group-average ERPs were an imperfect guide to individual detectability in both healthy adults and patients with MCS. In healthy adults, most paradigms elicited canonical responses, with SON being an exception. Yet a strong group-level N400 effect coexisted with substantially lower single-subject detectability, illustrating that robust group statistics do not guarantee reliable individual detection. In patients, the discrepancy ran in the opposite direction: grand averages often lacked canonical components, although some patients showed several ERPs resembling those of healthy controls. Variation in etiology, lesion profiles, and arousal may obscure responses present in only a subset of patients. Our findings demonstrate the importance of single-subject analysis, with healthy control detection rates providing a practical upper bound benchmark for interpreting patient results.

The highest single-subject classification accuracy was achieved in the oddball family of tasks, especially for MMN. Rutiku et al.^41^ reported similarly high within-subject detection performance in 15 participants using the Optimum-1 and local-global paradigms. In the local-global paradigm, Faugeras et al.^42^ reported the MMN in 7/8 healthy controls (87.5%).

In our MCS sample, MMN detection for the short deviant was 27%, which closely matches Fischer et al.^9^ (27%) and Faugeras et al.^42^ (32.1%). Some studies reported much higher rates in very small samples: de Tommaso et al.^43^ reported 100% MMN detection in 4 MCS patients, but such estimates are difficult to generalize. When visual scoring considered MMN responses to any deviant type, the rate increased to 65%. It is possible that some patients responded to one deviant but not to another due to anatomical differences in the source of the MMN suggesting that multifeature paradigms such as Optimum-1 may improve sensitivity in DOC. On the other hand, such paradigms raise a multiple-comparisons problem if no clear correction strategy is applied.

What does the lack of basic responses such as MMN in some patients tell us? In theory, the relative independence of the MMN from attention makes it a good marker of preserved sensory discrimination in DOC. Prior work has linked MMN presence to improved chances of awakening from coma^44^. At the same time, current clinical guidelines stress that sensitivity in DOC studies is often modest and that absence of MMN cannot be interpreted as evidence for absence of residual cognition^4,6^.

Oddball P300 effects were among the most reliable responses in the healthy group. This applies to both passive and active oddball tasks, and supports the idea that oddball P300 is one of the most reproducible ERP phenomena in cognitive neuroscience. Because of this reliability, oddball P300 is also a strong candidate for technical testing of EEG systems. If a device cannot detect the oddball effect in nearly all healthy participants, the signal-to-noise ratio is likely insufficient for more demanding clinical paradigms.

In our sample, active oddball P300 was present in around 31% of patients, which is consistent with previous reports. Steppacher et al.^10^ reported a similar proportion, with 11/37 MCS patients showing P300. Faugeras et al.^42^ reported a lower rate for the more demanding global P3b effect, 4/28 recordings (14.3%), which is expected because the global-local paradigm places stronger demands on temporal integration and task processing. Overall, this pattern supports the view that P300 is a useful marker of residual auditory-attentional processing in MCS patients.

N400 showed a clear dissociation between group-level strength and single-subject detectability. In healthy controls, the group-level effect size was large and similar to passive P300, but single-subject detection was clearly lower, with around two thirds or fewer participants classified as positive. This indicates greater interindividual variability in N400 morphology, timing, or topography than in oddball P300.

Our healthy control N400 detection rate of 69% fits well within the range reported previously. Across studies, single-subject detection in healthy participants has ranged from 17% to 88%, including 17% (2/12) and 50% (6/12) in two variants reported by Cruse et al.^26^, 57% (11/19) in Rohaut et al.^45^, 62% (48/79) in Kallionpää et al.^27^, and 88% (14/16) in Beukema et al.^16^. In patients, our detection rate was 3/26 (12%), which is low but still falls within the broad range of prior reports: 12.5% (1/8) in Beukema et al. (2016), 20.5% (8/39) in Steppacher et al. (2013), 21% (8/38) in Kotchoubey et al. (2005), 36% (5/14) in Rohaut et al. (2015), and 62.5% (5/8) in Formisano et al.^46^.

The breadth of these estimates suggests that N400 detectability depends not only on individual differences, but also on paradigm design. In particular, active N400 paradigms may improve detection in healthy participants^16,26,27^, but they also require preserved instruction comprehension and sustained attention, which limits their applicability in many MCS patients. From a clinical perspective, passive N400 paradigms do not appear to provide a highly sensitive marker of preserved language processing in MCS.

We used two tasks requiring command-following. In healthy volunteers, both performed well. Command-following inferred from active versus passive oddball P300 modulation reached 81%, and the semantic oddball task reached 92%. Our visual classification rate for semantic oddball was even higher than that for oddball-based command following, which suggests that semantic oddball may be a more efficient single-task option than running passive and active oddball back-to-back.

In active oddball tasks, a likely reason for the absence of an additional increase in P300 amplitude in some participants is reduced task engagement, since P3b can be strongly attenuated or abolished when attention is directed away from the relevant stimuli^47,48^. In our healthy sample, we could partially verify task engagement using reported counts. Most participants counted the deviants reasonably well, and only two showed large deviations from the expected number. Despite this, visual scoring detected P300 responses in every healthy participant in the oddball paradigms. Faugeras et al.^42^ also reported perfect detectability in healthy participants, including P3b in all 8 participants in the local-global paradigm.

Previous work using P300-based command-following paradigms in DOC also reported high detection rates in controls: Lulé et al.^49^ found 14/16 healthy controls positive (87.5%). Across the broader oddball command-following literature, detection rates in MCS patients vary depending on task design and comparison strategy. Schnakers et al.^20^ reported 9/14 MCS patients with task-related active modulation. Schnakers et al.^21^ reported 8/16 patients with enhanced P3 amplitude in active versus passive listening, and in the same study active counting of own-name occurrences versus passive listening identified 9/20 MCS patients. These rates are impressive given the task complexity, but other studies show that sensitivity can drop sharply with more demanding paradigms. Pokorny et al.^50^, using a dual-stream auditory attention task, reported successful classification in 8 out of 10 healthy controls but 0 out of 12 in MCS patients.

In our patient sample, we identified 2 patients with command following in the semantic oddball task. This is slightly lower than the rate reported in the only previous study using this paradigm in DOC, which reported 5/38 (Kotchoubey et al., 2005). However, we are cautious about interpreting these 2 positives as strong evidence of consciousness because neither patient showed any other significant effects, including MMN. This pattern raises the possibility of false positives.

The SON paradigm produced unexpectedly low detection rates. This was already evident at the group level, where the effect was not significant, and it translated into low single-subject detection rates in both healthy controls and MCS patients. This contrasts with prior studies. Sergent et al.^18^ reported 60% (9/15) and Schnakers et al.^20^ reported perfect detection rate of P300 to the patient’s own name 100% (12/12) single-subject detection in healthy volunteers. In MCS patients, some groups reported perfect detection of P300 to the patient’s own name: (6/6)^14^ and (14/14)^20^. Meanwhile, Sergent et al.^18^ reported that 50% (4/8) of MCS patients showing elevated positivity response to their own name compared with the other name. A likely explanation of our failure is that our implementation embedded the names in a conditioning design, which may have reduced the salience of the name stimulus. If the presentation of one’s own name becomes predictable because it is paired with a conditioned tone, the emotional or self-referential impact may be weakened. Given the low healthy-control detectability, the current SON implementation should be redesigned and revalidated before being considered for clinical use.

Considering all paradigms, only a minority of MCS patients showed detectable higher order ERP effects, while basic auditory change detection was present in about two thirds of the sample. Such low detection rates are striking in patients with established behavioral evidence of consciousness, many of whom could follow commands. Low detection in passive P300 and N400 paradigms further shows that this discrepancy extends beyond explicit command following. Even passive tasks may exceed patients’ available sensory or cognitive resources, while altered EEG responses after brain injury may make ERPs less consistent and harder to detect. A similar dissociation was reported by Bodien et al.^52^, who found no task-based fMRI or EEG response in 62% of patients with observable command following.

Our hierarchical approach was designed to probe progressively more complex processing, from automatic discrimination to semantic processing and command following. In the patient data, this hierarchy was present to some extent, but not perfectly. In most cases, lower-level responses such as MMN and passive or active oddball P300 were present in patients who also showed higher-order effects. Conversely, many patients with no MMN also showed no responses in any other paradigm. At the same time, there were exceptions. A few patients showed a semantic oddball or N400 effect without a detectable MMN or passive oddball response. In our sample, there were only three such cases with higher-order components present but without detected MMN. This means the hierarchy is informative but not absolute. Also, absence of MMN cannot be treated as strong evidence for absence of preserved mid-level sensory processing in a specific patient.

Our approach is conceptually similar to earlier efforts to probe multiple cognitive capacities in a single EEG session. Sergent et al.^18^ tested several dimensions of cognition in a 1.5 h protocol. They observed heterogeneous profiles, including cases where higher-level effects appeared without lower-level effects. The advantage of a hierarchical battery is that it gives a richer cognitive profile than a single paradigm. The main weakness is that long batteries can increase fatigue and state fluctuations.

Regarding the suitability of our current battery for clinical practice, some paradigms should be improved. In addition to the SON paradigm, the passive N400 paradigm also appears substandard for single-subject clinical use. It may be replaced by a more reproducible language task. A simpler contrast such as words versus pseudowords may yield better single-subject sensitivity. Supporting this, Ben Salah et al.^53^ reported that only 1 healthy control out of 13 did not show a significant ERP effect (N400 or P600).

Nearly all healthy participants showed significant effects in most paradigms with at least one of the classification methods, but the methods differed in sensitivity. ANOVA and cluster-based permutation tests yielded lower classification rates overall, which makes them more conservative than visual scoring and machine learning in our dataset. Kallionpää et al.^27^ also found that cluster permutation tests were not the most sensitive approach for N400 detection in healthy subjects.

Visual scoring was the most balanced method in our data. It gave very high rates in paradigms with strong expected effects and low rates in the SON paradigm, where even the group-level effect was absent. Initial agreement between the two scorers was above 80% for all paradigms except SON, which is acceptable, but not excellent. Although disagreements were resolved in consensus meetings, this process is time-consuming and still potentially vulnerable to subjective bias.

Machine learning was the second-best method overall and showed higher detection rates than visual scoring for active-versus-passive command following in both controls and patients. This fits the broader BCI literature, where ML is often preferred for single-subject decoding because it can account for individual variability in the spatial and temporal localization of the effect. At the same time, some of our ML-positive findings may reflect false positives. For example, ML yielded relatively high SON positivity in patients despite low visual detection and weak group-level effects. These could be true spatially distributed effects, but they could also reflect overfitting. A further caveat concerns single-subject SON contrasts. Within each participant, SON and OFN are acoustically different words preceded by different conditioned tones, so a reliable single-subject difference can arise from stimulus-specific auditory responses rather than self-relevance; counterbalancing removes such differences only at the group level. Consistent with this, the SON-OFN difference at Pz was positive in only 7 of 17 visually SON-positive controls and 19 of 35 ML-positive controls, whereas semantic oddball positives showed the expected polarity in nearly all cases (43 of 44 visually positive and 42 of 42 ML-positive controls). Single-subject SON detections by machine learning, therefore, should not be taken as good evidence of self-name recognition without directional criteria or an acoustic control.

Our findings align with comparative studies. Herrera-Diaz et al.^54^ found higher sensitivity for visual scoring than permutation-based methods in a multifeature MMN variant, at least for frequency and intensity deviants, while duration deviants reached 100% sensitivity with both methods. Gabriel et al.^29^ reported MMN in 20/27 conscious subjects by visual analysis and 17/27 with a simple predefined-window t test, which is conceptually close to our ANOVA approach. Overall, no single method is optimal for all paradigms, and a combined strategy may be preferable in clinical settings.

ERP presence did not predict 6-month outcome. The small sample (14 patients regained consciousness, 8 did not, and 4 died) limited power to detect meaningful associations. Only a few studies have formally predicted outcome using ERP variables alone^8^, and larger studies have also found that P300 is not a reliable predictor^55^. Our findings extend this caution to N400 and semantic oddball. This negative finding does not mean ERPs are clinically irrelevant, but it does suggest that single binary ERP indicators, at least in a sample of this size, have limited standalone prognostic value.

### Limitations

We did not explore the full multiverse of possible analyses, including time windows, and channels for every method. There are many other reasonable analytical choices, and some of our decisions were necessarily arbitrary, even though they were guided by prior ERP and DOC work.

Additional limitations include the relatively small patient sample for prognostic modeling, the fixed order of paradigms within each group, and the use of different EEG systems in controls and patients. Although we harmonized channels and preprocessing, some residual system-related variance is unavoidable. Finally, reducing each paradigm to a binary present versus absent classification likely discarded useful information about effect size, latency, and topography, but was necessary to keep the analysis manageable.

## Data Availability

All data produced in the present study are available upon reasonable request to the authors

## Data availability

Raw EEG data will be shared in BIDS format upon publication. The richness of data recorded in 6 common ERP paradigms (auditory) may serve for benchmarks and extend the currently available ERP CORE dataset^56^ that used primarily visual stimuli.

## Supplementary materials

**Supplementary Figure S1.**
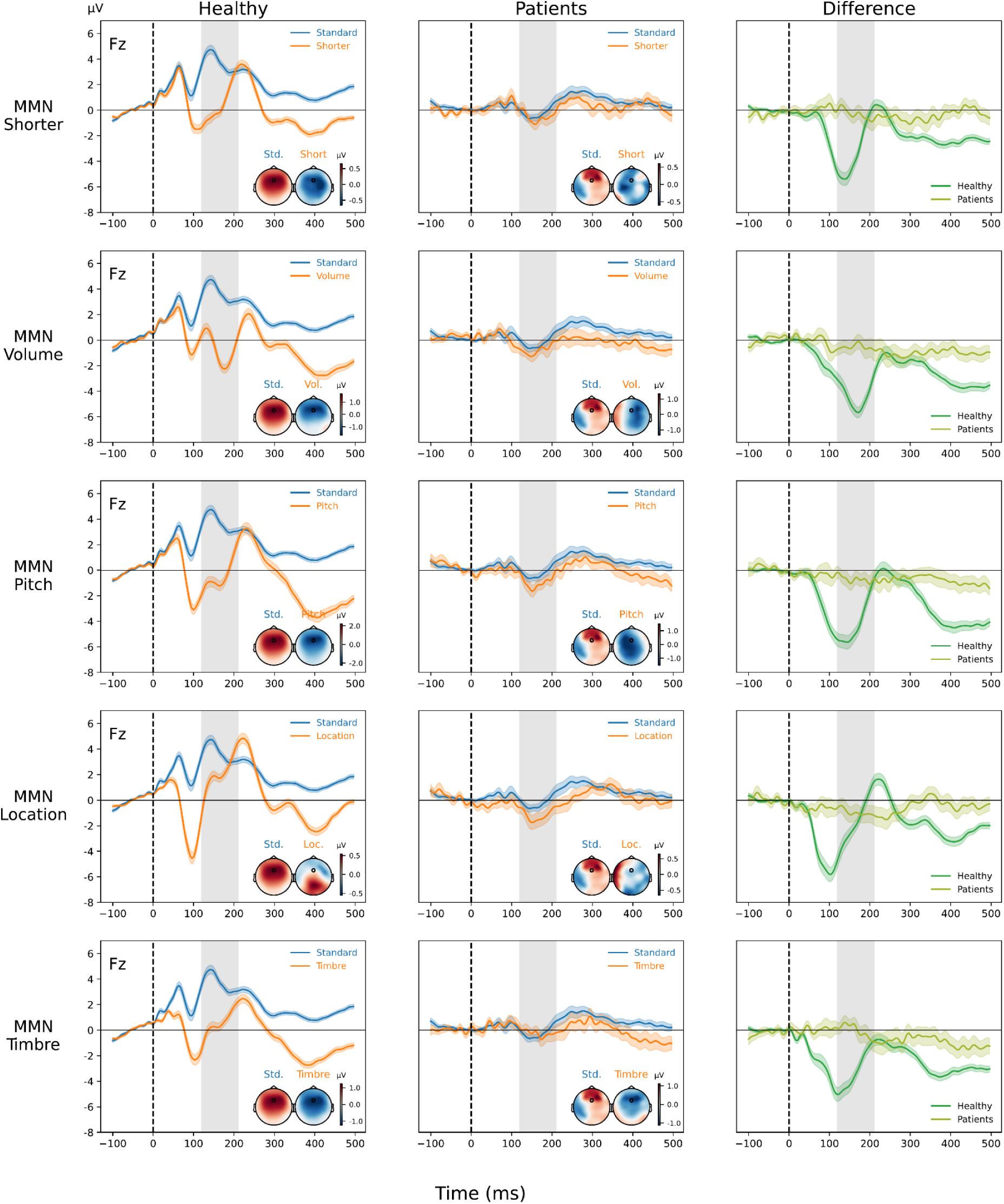
: Grand-average ERPs for all five mismatch negativity (MMN) deviant types. Responses at Fz are shown for healthy controls (n = 48; left column) and patients with minimally conscious state (n = 26; middle column). Rows show duration (“Shorter”), intensity (“Volume”), pitch, location, and timbre deviants. Blue and orange traces represent responses to standards and deviants, respectively. The right column shows deviant-minus-standard difference waves for healthy controls (green) and patients (yellow-green). Coloured bands indicate SEM. Dashed vertical lines mark stimulus onset, and grey shading indicates the 120–210 ms analysis window. Insets show scalp distributions of mean amplitude within this window, with Fz marked. Amplitudes are expressed in µV, with positive values plotted upwards.

